# Video-Based Pelvic Floor Muscle Therapy for Adult Patients with Pelvic Floor Disorders: A Protocol for a Prospective Single-Arm Interventional Pilot and Feasibility Study

**DOI:** 10.1101/2025.08.01.25332599

**Authors:** Samantha M. Linhares, Madeline L. D’Aquila, Kurtz S. Schultz, Anne K. Mongiu

**Author notes:** These authors contributed equally to this work.

## Abstract

The number of patients who suffer from pelvic floor disorders increases with age and can have a significant impact on quality of life. The first-line treatment for these different disorders includes pelvic floor rehabilitation. However, there are high rates of non-compliance with completing the recommended duration of treatment due to delays in appointments and time constraints. Thus, the primary goal of this study is to evaluate the feasibility and acceptability of an online 8-week video-based pelvic floor muscle therapy program. A secondary goal is determining whether the treatment can improve quality of life and symptoms. This study is a registry-based pilot single-arm prospective trial (NCT06689891: Video-Based Pelvic Floor Muscle Therapy). Eligible participants will be offered the online program as an alternative to in-person pelvic floor rehabilitation. Primary timepoints include a pre-intervention in-person visit with a licensed pelvic floor therapist and the 8-week video-based pelvic floor muscle therapy program. There will be a midpoint evaluation followed by a post-intervention visit with the same pelvic floor therapist, where participants will be graded on their ability to complete the various exercises to assess efficacy. A survey assessing the online-based program’s usability will be conducted post-intervention. Patient-reported outcome measures, including quality-of-life and symptom changes, will be collected pre-, mid-, and post-intervention. As this is a pilot trial, the goal is to establish the acceptability and feasibility of a video-based pelvic floor muscle therapy program as an alternative to in-person treatment.

## INTRODUCTION

Pelvic floor disorders, which include pelvic organ prolapse, urinary incontinence, and anorectal dysfunction, affect up to 25-32% of women during their lifetime.^1,2^ These disorders can be debilitating and will affect quality of life (QoL) for up to 50% of patients.^3^ Pelvic floor rehabilitation (PFR) is the first-line treatment for patients with pelvic floor disorders and is part of the management algorithm for other disease processes, such as low anterior resection syndrome (LARS).^4–6^ The exercises commonly used in PFR include pelvic floor muscle therapy (PFMT) with biofeedback training, rectal balloon training, or electrical stimulation.^7–9^ Together, these techniques improve the strength, tone and endurance of the pelvic floor musculature and sphincter complex.^8^ They have decreased symptoms ranging between 50-80% for fecal incontinence.^10,11^ A meta-analysis of 18 trials utilizing PFMT for urinary incontinence found an improvement in symptoms in all studies.^12^

Despite PFMT being shown to be efficacious for numerous different pelvic floor dysfunction disorders, less than 50% of patients referred are compliant with the recommended duration of treatment.^13,14^ A study of 180 patients referred for PFR found that 34% of patients did not attend a single session, and only 29% completed the entire duration.^15^ Previous studies found that inconvenience, cost, lack of knowledge, and discomfort around the concept of PFMT are all reasons why patients may have poor compliance.^16–18^ Patients have shown a positive reception to telehealth-based PFMT visits.^19,20^ It is unknown if a hybrid approach to PFMT, including both in-person and video-based training, may help alleviate these barriers while providing the same adequate amount of guidance, improvement in symptoms, and QoL for patients. Thus, this study aims to evaluate the feasibility and acceptability of an online 8-week video-based PFMT for pelvic floor disorders. We hypothesize that an online video-based PFMT program will be a feasible alternative to in-person pelvic floor muscle therapy for patients.

## MATERIALS AND METHODS

This study is a registry-based feasibility and acceptability prospective single-arm trial (NCT06689891: Video-Based Pelvic Floor Muscle Therapy).^21^ The protocol has received approval from the Yale University Institutional Review Board (#2000037163). This study aims to evaluate the feasibility and acceptability of an online video-based PFMT program, a novel delivery model for a proven first-line treatment for pelvic floor disorders. This will be a hybrid design with two in-person pelvic floor therapy sessions at a tertiary academic hospital and the participant’s permanent residence in the United States. The schedule of enrollment, interventions, and assessments is shown in Figure 1. Study enrollment began on June 6, 2025. Enrollment is ongoing, and the recruitment period has not yet ended. We estimate participant recruitment will be completed by December 2025. Data collection will be completed by March 2026. We expect to have preliminary results by April 2026.

**Figure 1.**
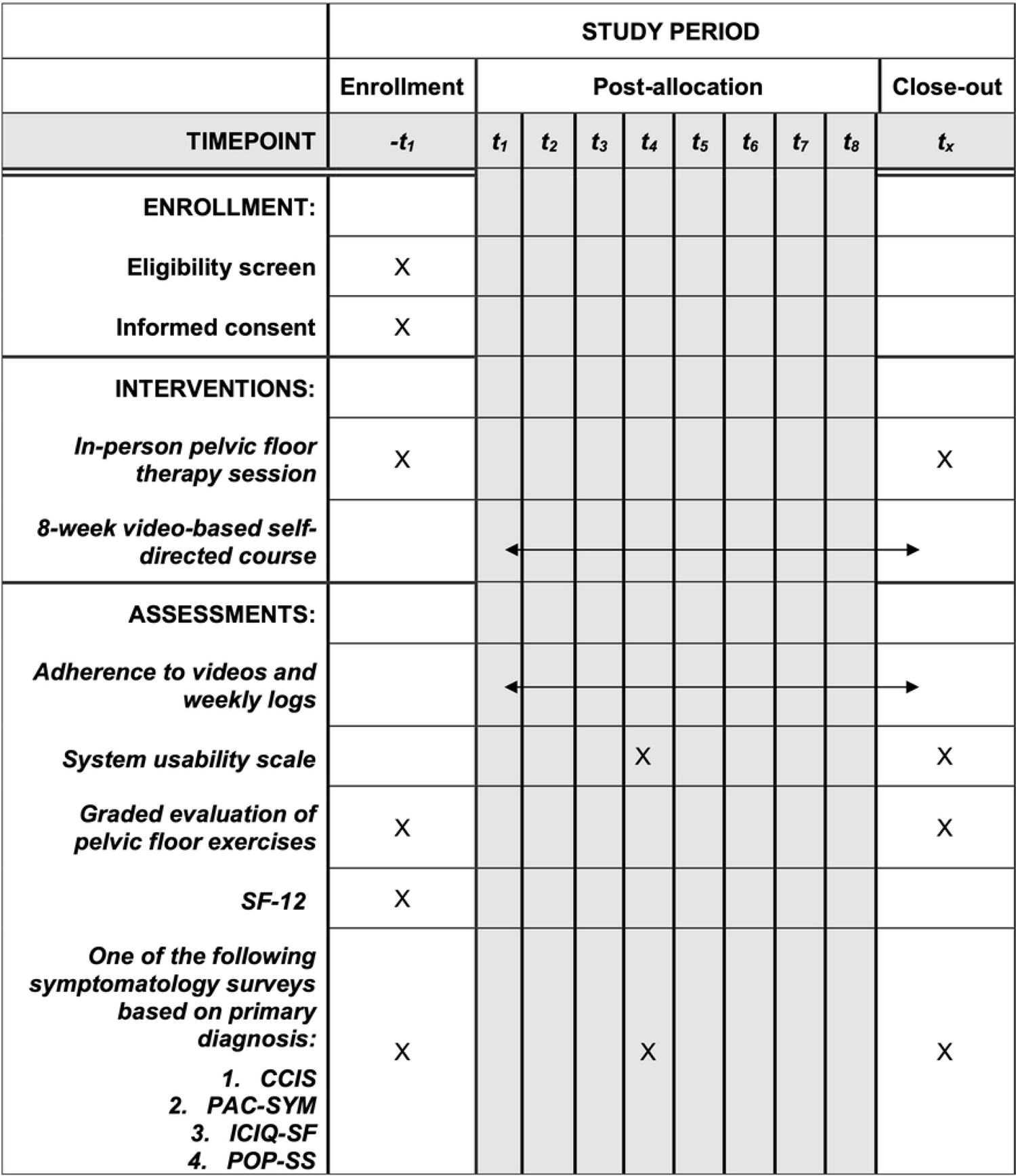
SPIRIT schedule of enrollment, interventions, and assessments.

### Sample size

As this is a pilot study, we will aim to recruit 20 adult participants, which is within the range typical of other pilot studies to test feasibility and acceptability.^22,23^ Study sites include outpatient urogynecology clinic and colorectal surgery clinic. Twenty English-speaking individuals at least 18 years of age with a diagnosis of urinary incontinence, pelvic organ prolapse, pelvic pain, fecal incontinence, or constipation will be recruited during their urogynecology or colorectal surgery clinic visit for one of these conditions (Table 1). Exclusion criteria include patients who cannot speak or read English, those who do not have reliable access to the internet, and those who do not know how to navigate the internet. Researchers will recruit patients through follow-up emails or phone calls to patients who have expressed initial interest in the study to their primary healthcare provider.

**Table 1.**
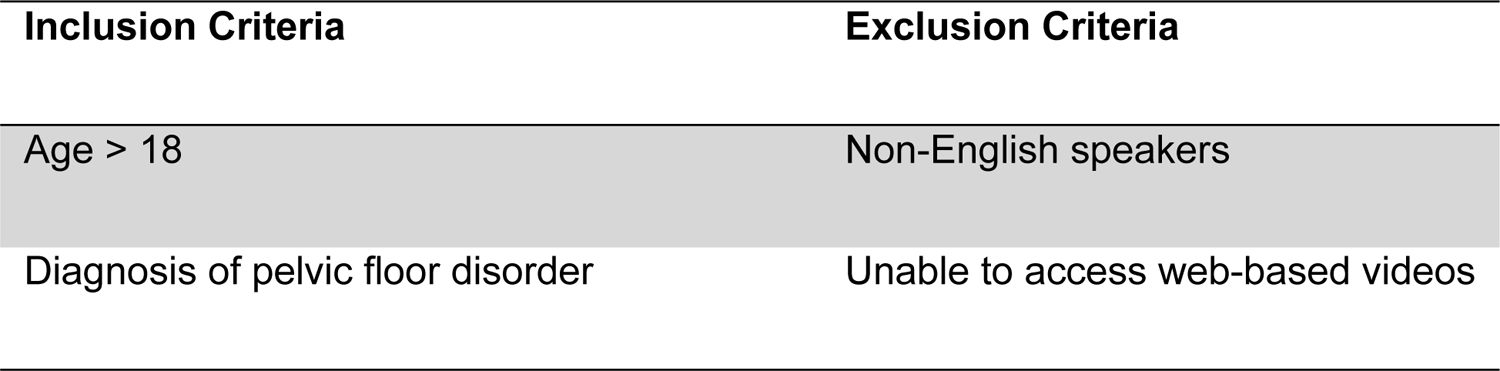
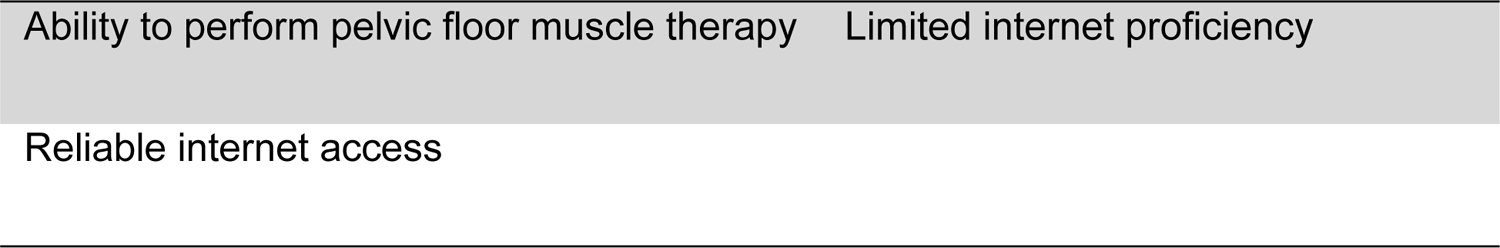
Inclusion and exclusion criteria.

### Intervention

The baseline study visit will be a standard of care visit with an in-person pelvic floor therapist. This visit will introduce the concepts important to pelvic floor exercises and assess the participant’s baseline function with graded evaluation of a standard set of pelvic floor exercises, which is discussed further in the efficacy outcome below. Participants will then complete eight weeks of online video-based PFMT. The videos have four sections: introduction followed by beginner, intermediate and advanced exercises demonstrations. Each set of exercises is a 30-minute video. Participants are recommended to engage with the program for 20-35 minutes at least three days a week. Participants will be sent a weekly log to record the number of days they exercised. There will be mid-point feedback at 4 weeks, where participants will complete the SUS and repeat the initial surveys to assess QoL and symptomatology. After the video-based PFMT program is completed, participants will have a second in-person session with the same PFPT for follow-up and repeat graded assessment of the PFMT exercises. They will complete the System Usability Scale (SUS), QoL, and appropriate symptomatology survey again within two weeks of the final PFPT appointment (Figure 2).

**Figure 2.**
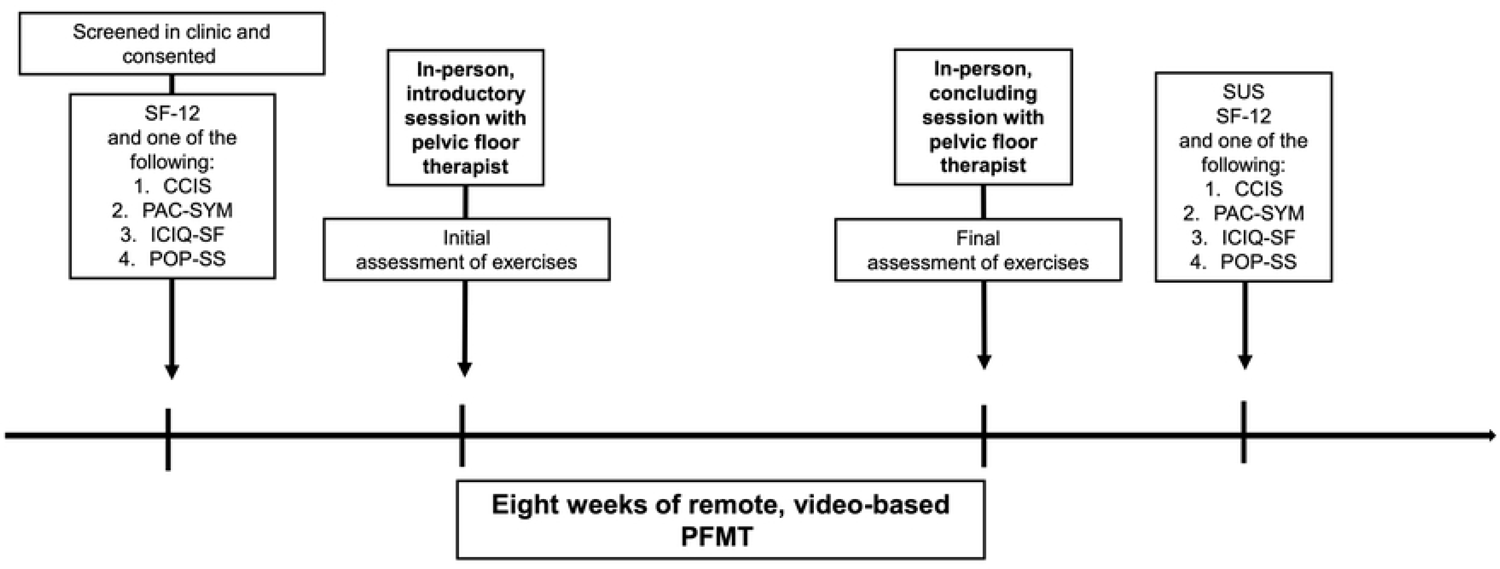
Study design for a proposed feasibility trial of using video-based pelvic floor muscle therapy (PFMT) intervention for patients. At the initial clinic visit where patients are eligible for receiving PFMT, they will be offered to enroll in video-based PFMT. If they are consented, they will asked to complete the Short Form Health Survey (SF-12), and one of the four following symptom-specific surveys: 1) Cleveland Clinic Incontinence Score (CCIS), 2) Patient Assessment of Constipation Symptoms (PAC-SYM), 3) International Consensus on Incontinence Questionnaire Short Form (ICIQ-SF) or 4) Pelvic Organ Prolapse Symptom Score (POP-SS) which they will complete again after the 8 weeks of video-based PFPT. After conclusion of the 8 weeks, they will also complete the System Usability Survey (SUS) to evaluate for ease of use of the web-based application. They will meet with a pelvic floor therapist prior to initiation of the video-based PFMT and again at the conclusion with an assessment of exercise competency preoperatively and postoperatively.

### Relevant concomitant care or prevention

During this study period, patients will be asked to abstain from participating in additional supervised in-person pelvic floor therapy sessions. Patients may choose at any point to opt out of the study and pursue the standard of care with in-person supervised pelvic floor therapy for patient preference or worsening symptoms. Patient symptomatology and QoL will be assessed at the midpoint feedback and patients will be notified if their symptoms are improved or worsened. They will then be offered the opportunity to complete additional recommended number of sessions in person.

### Primary outcomes

#### Feasibility

The primary outcome measure for feasibility will be patient adherence to the 8-week online video base PFMT program, which will be measured via weekly logs and completion of all videos. We will aim for patients to have maintained at least 75% adherence to recommended weekly exercise amount, as is common among exercise or physical activity intervention studies.^24–26^

#### Acceptability

The SUS is the primary outcome measure for acceptability. This 10-item questionnaire was initially developed in 1986 to capture a user’s evaluation of an online system’s usability with a 10-item questionnaire.^27,28^ It is the most widely used questionnaire to evaluate health technology interfaces and has robust data for validation.^29,30^ The SUS will be administered at the midpoint and completion of the study to determine the ease of independently following the videos and exercises.

### Secondary outcomes

Secondary outcomes will include efficacy and

#### Efficacy

The outcome measure for efficacy is a pelvic floor therapist’s graded evaluation of exercises. The graded assessment is based on evaluating a set of standard of care pelvic floor exercises, ranging from easy to more difficult, as has been used in other pelvic floor therapy studies.^31^ Each exercise will be given a score of one to three, with one indicating that the participant is unable to complete the exercise without assistance, two indicating the participant can partially complete the exercise independently, and a three indicating the participant can complete the exercise independently. A total of five exercises of varying difficulty will be assessed.

#### Quality of life

Quality of life changes are assessed with the Short Form-12 (SF-12), along with changes in symptomatology based on one of the four appropriate symptom-focused surveys: 1) Cleveland Clinic Incontinence Score (CCIS), 2) Patient Assessment of Constipation Symptoms (PAC-SYM), 3) International Consultation on Incontinence Questionnaire Short Form (ICIQ-SF), and 4) Pelvic Organ Prolapse Symptom Score (POP-SS)

The SF-12 is a 12-item questionnaire to assess quality of life in the physical and mental domains. It is an abbreviated form of the longer SF-36, which is a 36-item questionnaire. The SF-12 has been tested with good reliability and validity compared to the SF-36, and we have opted to use this shorter survey to reduce the burden on participants.^32^ The CCIS is one of the most common instruments for assessing fecal incontinence.^33^ This is a five-item questionnaire designed to assess patient symptomatology related to incontinence in terms of type of leakage (i.e., solid versus liquid versus flatus), the necessity to wear hygiene pads, and the impact on lifestyle. Compared to several commonly used incontinence scales, this questionnaire is equally validated and more succinct.^34^ The PAC-SYM is a 12-item questionnaire developed using patient perspectives to assess constipation on three symptom subscales: abdominal, rectal, and stool.^35^ It has been widely validated and is one of the most common assessments for constipation.^36^ The ICIQ-SF is a four-item questionnaire that assesses the frequency, severity, and impact on QoL of urinary incontinence and is the most commonly used questionnaire worldwide^37^ and is endorsed by multiple societies, including the American Society of Colon and Rectal Surgeons and the American Urogynecologic Society.^36^ The POP-SS is a seven-item questionnaire developed to assess the severity of pelvic organ prolapse symptoms and has been extensively validated^38,39^ (Table 2).

**Table 2.**
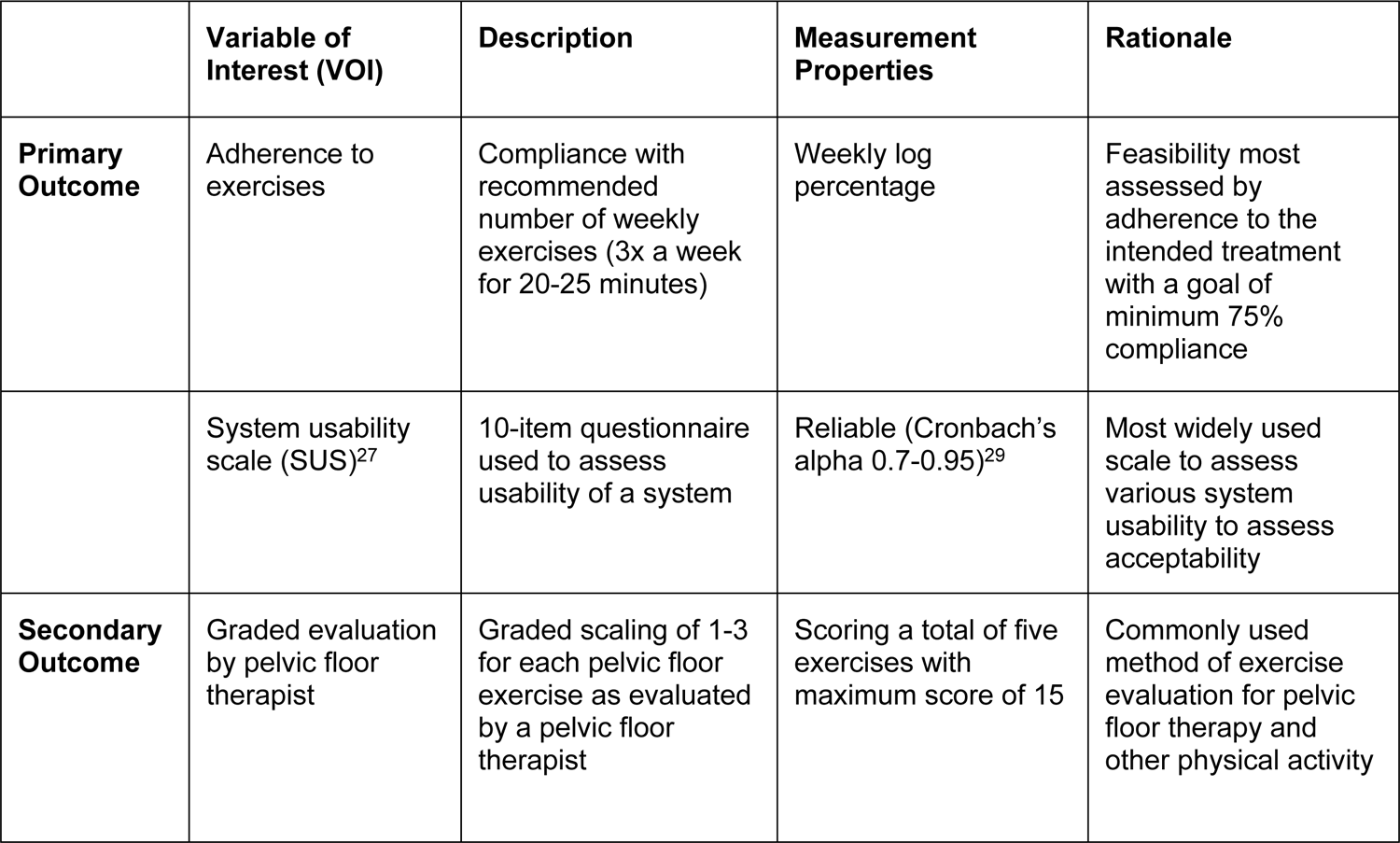
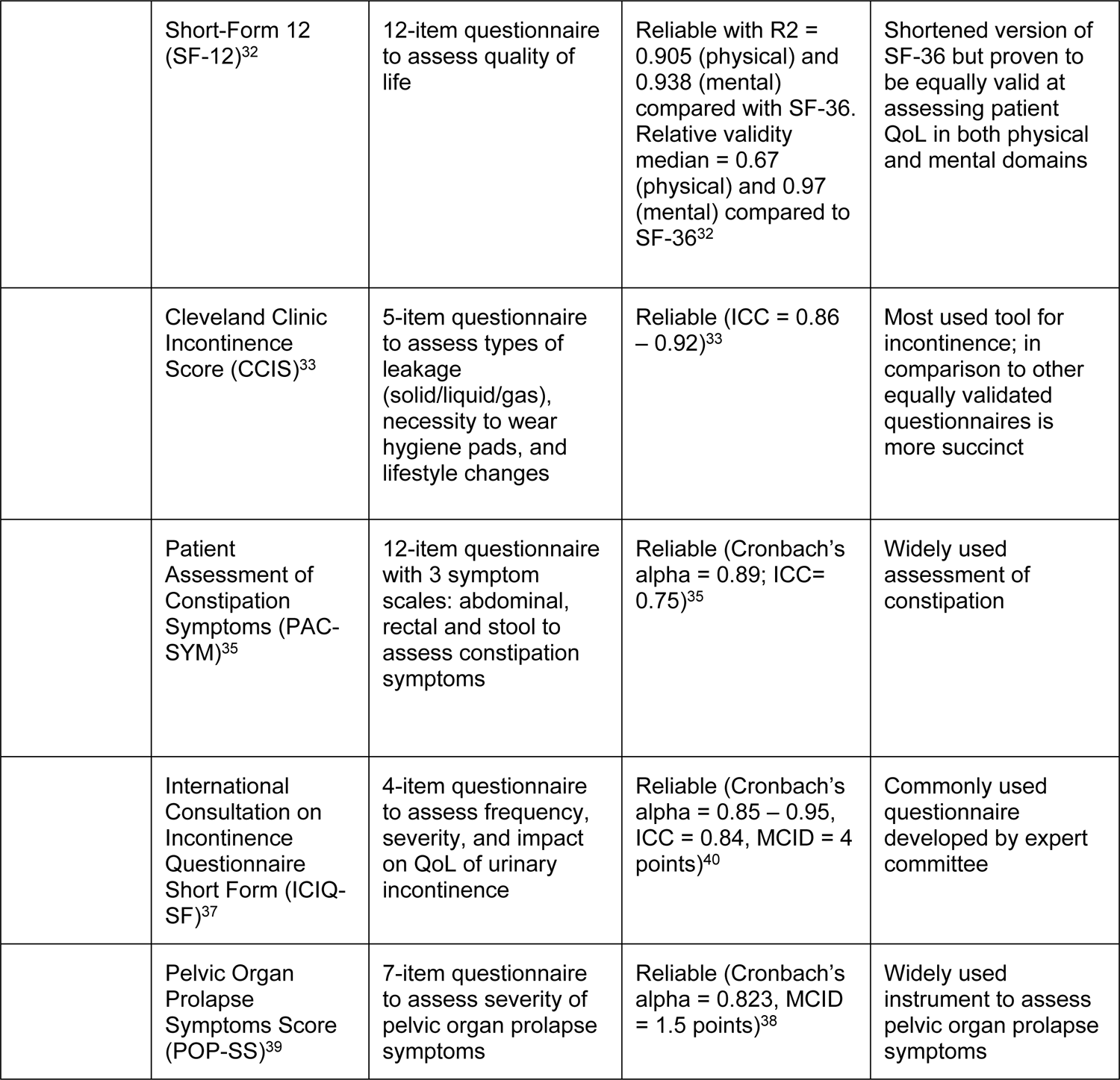
List of primary and secondary outcomes.

### Statistical analysis

An intention-to-treat approach will be used for all analyses. Normality will be evaluated, and non-parametric statistical tests will be used, as appropriate. Patient demographics and clinical characteristics, including age, sex, race, ethnicity, and indication for pelvic floor referral, will be summarized as descriptive statistics. The primary outcome of feasibility will be determined based on a minimum of 75% patient adherence to the program, which will be calculated based on the number of days completed per weekly log and completion of all the videos. Acceptability will be evaluated using the SUS scores at the midpoint and post-intervention. To evaluate the efficacy outcome, we will use the Wilcoxon signed-rank test to compare the graded evaluation scores by the pelvic floor therapist pre- and post-intervention. The other secondary outcomes will be treated as repeated measures and evaluated in a linear mixed effects model using the three timepoints (pre-intervention, midpoint, and post-intervention).

### Data management plan

The survey information will be either researcher-administered or independently completed by the study participant using an institution-licensed REDCap platform. If there are technical difficulties, a paper-based version of the assessment tool will be substituted and inputted into the system later. Demographic and clinical data will be extracted from the electronic medical records (e.g., Epic® chart reviews) and will be securely stored in the HIPAA-compliant REDCap database. The assessment by the pelvic floor therapist will also be securely input into the REDCap database. The full protocol, datasets, and statistical code used during this study will be made available from the corresponding author on reasonable request.

### Monitoring

The study is deemed minimal risk as patients can perform on their own and would not increase the magnitude of harm or discomfort than those ordinarily encountered in routine physical exams. The exercises as part of the program are not outside the scope of the standard of care. No interim analyses are planned for this study because the intervention is low-risk, and the primary purpose of this pilot study is feasibility not efficacy.

Oversight of trial safety and data integrity will be by the study team and internal institutional monitoring mechanisms at least monthly. In the unlikely event that an adverse event occurs, a written report will be submitted within five calendar days of the Principal Investigator becoming aware of the event to the IRB and appropriate funding and regulatory agencies. The study team and sponsor will review the data and determine if modification or early termination is warranted.

### Ethics and dissemination

The protocol has received approval from the Yale University Institutional Review Board (#2000037163). Authorized research study staff will obtain written informed consent from participants prior to starting the study. Important protocol changes will be communicated to participants, study staff, and the IRB by the study investigator.

Data will be securely stored in an institutional HIPPA protected online database with a built-in auditing system. Only authorized study staff will have access to the data. Study results will be submitted for presentation and publication at future national meetings and in surgical journals. Authorship criteria will be defined by the International Committee of Medical Journal Editors. Study investigators do not have any conflicts of interest to declare.

## DISCUSSION

This study protocol is designed to test the feasibility and acceptability of a web-accessible online eight-week PFMT program as an alternative delivery method to the standard of care for the treatment of PFD. PFD is a constellation of disorders that affects nearly one-third of the female population, with a notable impact on QoL.^3^ Despite PFMT being a first-line treatment for these disorders, many barriers are in place that prohibits patients from completing the recommended number of sessions or even attending one session. It is not known whether a hybrid, online video-based PFMT program would be a feasible and safe alternative to in-person PFR.

Some potential pitfalls to this project include a small sample size with multiple indications for PFR, which may increase variability in determining acceptability. To minimize this, we will group results into two broad indications: urogynecology and colorectal. Another pitfall is loss to follow-up and lack of adherence during the self-directed portion. To mitigate this, we will incentivize participants to complete the study by giving them $50 gift cards after the final in-person assessment. We will also provide weekly log check-ins to remind participants to continue exercising.

The results of this study have the potential to be transformative for individuals referred for PFR by establishing a novel healthcare delivery model for the treatment of pelvic floor disorders. This pilot trial will provide preliminary data for a larger, multicenter study to evaluate the use of this delivery model as a potential alternative or supplement to in-person pelvic floor therapy. This study’s delivery platform could also be adopted and assessed for other diseases and conditions, such as adapting successful prehabilitation programs to an online delivery model for patients prior to surgery.^41,42^ Online alternatives continue to be on the rise to provide patients with additional flexibility in an increasingly busy world. Development of a web-accessible video-based PFMT program would target an unmet need, benefit individuals suffering from a variety of PFD, and pave the way for other alternatives in care.

## SUPPORTING INFORMATION

**S1 Checklist**. SPIRIT checklist.

**S2 Protocol**. Clinical study protocol approved by the institutional review board.

## Data Availability

No datasets were generated or analysed during the current study. All relevant data from this study will be made available upon study completion.

## REFERENCES

1. Kenne KA, Wendt L, Brooks Jackson J. Prevalence of pelvic floor disorders in adult women being seen in a primary care setting and associated risk factors. Scientific Reports. 2022/06/14 2022;12(1):9878. doi:10.1038/s41598-022-13501-w

2. Nygaard I, Barber MD, Burgio KL, et al. Prevalence of symptomatic pelvic floor disorders in US women. Jama. 2008;300(11):1311–1316.

3. Palmieri S, De Bastiani SS, Degliuomini R, et al. Prevalence and severity of pelvic floor disorders in pregnant and postpartum women. Int J Gynaecol Obstet. Aug 2022;158(2):346–351. doi:10.1002/ijgo.14019

4. Linhares SM, Schultz KS, Coppersmith NA, et al. Association Between Chemotherapy-Induced Peripheral Neuropathy and Low Anterior Resection Syndrome. Cancers. 2024;16(21):3578.

5. Fick CN, Linhares SM, Schultz KS, et al. Coding the issue: low anterior resection syndrome following rectal cancer treatment. Opinion. Frontiers in Surgery. 2024-November-18 2024;Volume 11 - 2024doi:10.3389/fsurg.2024.1503410

6. Coppersmith NA, Schultz KS, Esposito AC, et al. Colorectal surgeon practice patterns of low anterior resection syndrome after rectal cancer treatment. Supportive Care in Cancer. 2025/02/24 2025;33(3):218. doi:10.1007/s00520-025-09290-3

7. Dulskas A, Smolskas E, Kildusiene I, Samalavicius NE. Treatment possibilities for low anterior resection syndrome: a review of the literature. International journal of colorectal disease. 2018;33:251–260.

8. Scott KM. Pelvic Floor Rehabilitation in the Treatment of Fecal Incontinence. Clin Colon Rectal Surg. 2014/09/24 2014;27(03):99–105. doi:10.1055/s-0034-1384662

9. van der Heijden J, Kalkdijk-Dijkstra A, Pierie J, et al. Pelvic floor rehabilitation after rectal cancer surgery: A multicenter randomized clinical trial (FORCE trial). LWW; 2022.

10. Damon H, Siproudhis L, Faucheron J-L, et al. Perineal retraining improves conservative treatment for faecal incontinence: a multicentre randomized study. Digestive and Liver Disease. 2014;46(3):237–242.

11. Norton C, Whitehead W, Bliss D, Harari D, Lang J. Management of fecal incontinence in adults. Neurourology and Urodynamics: Official Journal of the International Continence Society. 2010;29(1):199–206.

12. Dumoulin C, Hay-Smith J, Habée-Séguin GM, Mercier J. Pelvic floor muscle training versus no treatment, or inactive control treatments, for urinary incontinence in women: a short version Cochrane systematic review with meta-analysis. Neurourology and urodynamics. 2015;34(4):300–308.

13. Ross JH, Sinha A, Propst K, Ferrando CA. Adherence to Pelvic Floor Physical Therapy Referrals in Women With Fecal Incontinence. Female Pelvic Med Reconstr Surg. Mar 1 2022;28(3):e29–e33. doi:10.1097/SPV.0000000000001140

14. Woodburn KL, Tran MC, Casas-Puig V, Ninivaggio CS, Ferrando CA. Compliance with pelvic floor physical therapy in patients diagnosed with high-tone pelvic floor disorders. Urogynecology. 2021;27(2):94–97.

15. Shannon MB, Genereux M, Brincat C, et al. Attendance at Prescribed Pelvic Floor Physical Therapy in a Diverse, Urban Urogynecology Population. Pm r. Jun 2018;10(6):601–606. doi:10.1016/j.pmrj.2017.11.008

16. Dumoulin C, Hay-Smith J, Frawley H, et al. 2014 consensus statement on improving pelvic floor muscle training adherence: International Continence Society 2011 State-of-the-Science Seminar. Neurourology and urodynamics. 2015;34(7):600–605.

17. Washington BB, Raker CA, Sung VW. Barriers to pelvic floor physical therapy utilization for treatment of female urinary incontinence. American journal of obstetrics and gynecology. 2011;205(2):152. e1-152. e9.

18. Zoorob D, Higgins M, Swan K, Cummings J, Dominguez S, Carey E. Barriers to pelvic floor physical therapy regarding treatment of high-tone pelvic floor dysfunction. Urogynecology. 2017;23(6):444–448.

19. ter Haar CM, Class QA, Kobak WH, Pandya LK. Telehealth in a Pelvic Floor Physical Therapy Clinic: A Retrospective Cohort Study. Urogynecology. 9900:10.1097/SPV.0000000000001510. doi:10.1097/spv.0000000000001510

20. Aloul ZS, Schultz KS, Linhares SM, et al. Patient-Reported Barriers to Completing In-Person Pelvic Floor Physical Therapy Referrals. Oral presentation (Quick Shot) presented at: American Society of Colon & Rectal Surgeons Annual Scientific Meeting; 2025/05/11 San Diego, CA.

21. Utilizing Remote, Video-Based Pelvic Floor Muscle Therapy for the Management of Low Anterior Resection Syndrome in Rectal Cancer Patients: A Feasibility Trial. National Library of Medicine (US). https://clinicaltrials.gov/study/NCT06689891

22. Billingham SA, Whitehead AL, Julious SA. An audit of sample sizes for pilot and feasibility trials being undertaken in the United Kingdom registered in the United Kingdom Clinical Research Network database. BMC Med Res Methodol. Aug 20 2013;13:104. doi:10.1186/1471-2288-13-104

23. Teresi JA, Yu X, Stewart AL, Hays RD. Guidelines for Designing and Evaluating Feasibility Pilot Studies. Med Care. Jan 1 2022;60(1):95–103. doi:10.1097/mlr.0000000000001664

24. Taylor TR, Makambi K, Sween J, Roltsch M, Adams-Campbell LL. The effect of a supervised exercise trial on exercise adherence among African American Men: a pilot study. J Natl Med Assoc. Jun 2011;103(6):488–91. doi:10.1016/s0027-9684(15)30362-x

25. Santos A, Braaten K, MacPherson M, et al. Rates of compliance and adherence to high-intensity interval training: a systematic review and Meta-analyses. Int J Behav Nutr Phys Act. Nov 21 2023;20(1):134. doi:10.1186/s12966-023-01535-w

26. MacDonald CS, Ried-Larsen M, Soleimani J, et al. A systematic review of adherence to physical activity interventions in individuals with type 2 diabetes. Diabetes Metab Res Rev. Nov 2021;37(8):e3444. doi:10.1002/dmrr.3444

27. Lewis JR. The system usability scale: past, present, and future. International Journal of Human–Computer Interaction. 2018;34(7):577–590.

28. Brooke J. SUS-A quick and dirty usability scale. Usability evaluation in industry. 1996;189(194):4–7.

29. Hyzy M, Bond R, Mulvenna M, et al. System usability scale benchmarking for digital health apps: meta-analysis. JMIR mHealth and uHealth. 2022;10(8):e37290.

30. Brooke J. SUS: a retrospective. Journal of usability studies. 2013;8(2)

31. Betschart C, Mol SE, Lütolf-Keller B, Fink D, Perucchini D, Scheiner D. Pelvic floor muscle training for urinary incontinence: a comparison of outcomes in premenopausal versus postmenopausal women. Female Pelvic Med Reconstr Surg. Jul-Aug 2013;19(4):219–24. doi:10.1097/SPV.0b013e31829950e5

32. Ware J, Jr., Kosinski M, Keller SD. A 12-Item Short-Form Health Survey: construction of scales and preliminary tests of reliability and validity. Med Care. Mar 1996;34(3):220–33. doi:10.1097/00005650-199603000-00003

33. Jorge MJ, Wexner SD. Etiology and management of fecal incontinence. Diseases of the colon & rectum. 1993;36(1):77–97.

34. Norderval S, Rydningen MB, Falk RS, Stordahl A, Johannessen HH. Strong agreement between interview-obtained and self-administered Wexner and St. Mark’s scores using a single questionnaire. Int Urogynecol J. Dec 2019;30(12):2101–2108. doi:10.1007/s00192-019-03945-6

35. L. Frank LKCFLTPM. Psychometric Validation of a Constipation Symptom Assessment Questionnaire. Scandinavian Journal of Gastroenterology. 1999/01/01 1999;34(9):870–877. doi:10.1080/003655299750025327

36. Bordeianou LG, Anger JT, Boutros M, et al. Measuring Pelvic Floor Disorder Symptoms Using Patient-Reported Instruments: Proceedings of the Consensus Meeting of the Pelvic Floor Consortium of the American Society of Colon and Rectal Surgeons, the International Continence Society, the American Urogynecologic Society, and the Society of Urodynamics, Female Pelvic Medicine and Urogenital Reconstruction. Female Pelvic Med Reconstr Surg. Jan/Feb 2020;26(1):1–15. doi:10.1097/spv.0000000000000817

37. Avery K, Donovan J, Peters TJ, Shaw C, Gotoh M, Abrams P. ICIQ: a brief and robust measure for evaluating the symptoms and impact of urinary incontinence. Neurourology and Urodynamics: Official Journal of the International Continence Society. 2004;23(4):322–330.

38. Hagen S, Glazener C, Cook J, Herbison P, Toozs-Hobson P. Further properties of the pelvic organ prolapse symptom score: minimally important change and test-retest reliability. Neurourol Urodyn. 2010;29(6):1055–1056.

39. Hagen S, Glazener C, Sinclair L, Stark D, Bugge C. Psychometric properties of the pelvic organ prolapse symptom score. Bjog. Jan 2009;116(1):25–31. doi:10.1111/j.1471-0528.2008.01903.x

40. Hajebrahimi S, Nourizadeh D, Hamedani R, Pezeshki MZ. Validity and reliability of the International Consultation on Incontinence Questionnaire-Urinary Incontinence Short Form and its correlation with urodynamic findings. Urol J. Fall 2012;9(4):685–90.

41. Molenaar CJL, Minnella EM, Coca-Martinez M, et al. Effect of multimodal prehabilitation on reducing postoperative complications and enhancing functional capacity following colorectal cancer surgery: the PREHAB randomized clinical trial. JAMA surgery. 2023;158(6):572–581.

42. Coppersmith NA, Schultz KS, Esposito AC, et al. Pelvic Floor Physical Therapy Prehabilitation (PrePFPT) for the prevention of low anterior resection syndrome. Surgical Oncology Insight. 2024/12/01/ 2024;1(4):100097. 10.1016/j.soi.2024.100097

